# Performance of Generative Pretrained Transformer on the National Medical Licensing Examination in Japan

**DOI:** 10.1101/2023.04.17.23288603

**Authors:** Yudai Tanaka, Takuto Nakata, Ko Aiga, Takahide Etani, Ryota Muramatsu, Shun Katagiri, Hiroyuki Kawai, Fumiya Higashino, Masahiro Enomoto, Masao Noda, Mitsuhiro Kometani, Masayuki Takamura, Takashi Yoneda, Hiroaki Kakizaki, Akihiro Nomura

## Abstract

The remarkable performance of ChatGPT, launched in November 2022, has significantly impacted the field of natural language processing, inspiring the application of large language models as supportive tools in clinical practice and research worldwide. Although ChatGPT recently scored high on the United States Medical Licensing Examination, its performance on medical licensing examinations of other nations, especially non-English speaking nations, has not been sufficiently evaluated. This study assessed ChatGPT’s performance on the National Medical Licensing Examination (NMLE) in Japan and compared it with the actual minimal passing rate for this exam. In particular, the performances of both the GPT-3.5 and GPT-4 models were considered for the comparative analysis. We initially used a model and prompt tuning set of 290 questions without image data from the previous 116^th^ NMLE (held in February 2022) to maximize the performance for delivering correct answers and explanations of the questions. Thereafter, we tested the performance of the best ChatGPT model (GPT-4) with tuned prompts on a dataset of 262 questions without images from the latest 117^th^ NMLE (held in February 2023). The best model with the tuned prompts scored 82.7% for the essential questions and 77.2% for the basic and clinical questions, both of which sufficed the minimum passing rates of 80.0% and 74.6%, respectively. Simultaneously, we identified the three major factors contributing to the generation of the incorrect answers—insufficient medical knowledge, information on Japan-specific medical system and guidelines, and mathematical errors. In conclusion, GPT-4 powered ChatGPT with our optimally tuned prompts achieved a minimum passing rate in the latest 117^th^ NMLE in Japan. Although we express strong concerns regarding the use of the current ChatGPT for medical purposes so far, these artificial intelligence models may soon have the potential to serve as one of the best “sidekicks” for solving medical and healthcare problems.

**Author summary:** ChatGPT’s remarkable performance has inspired the use of large language models as supportive tools in clinical practice and research. Although it scored well in the US Medical Licensing Examination, its effectiveness in relevant examinations of non-English speaking countries remain unexplored. This study assessed the performance of ChatGPT with GPT-3.5 and GPT-4 models in Japan’s National Medical Licensing Examination (NMLE). Initially, we used a tuning set of 290 questions from the 116th NMLE, and then the GPT-4 model with tuned prompts was tested on 262 questions from the 117th NMLE. The model scored 82.7% for essential and 77.2% for basic and clinical questions, surpassing the minimum passing rates. Incorrect answers were attributed to insufficient medical knowledge, Japan-specific medical system information, and mathematical errors. In conclusion, GPT-4 powered ChatGPT achieved a minimum passing rate and might have the potential for a valuable tool for fulfilling the needs of medical and healthcare fields.

## Introduction

In recent decades, artificial intelligence (AI) algorithms have been widely applied in medical and healthcare fields [1]. Currently, the AI algorithms available for clinical applications have been developed using previous rule-based methods as well as recent machine learning (ML) methods including its subfield of deep learning, promoted by the continually increasing availability of computer resources and vast amount of medical data [2]. Consequently, these medical AI products have been implemented to obtain targeted outputs such as the *prediction* of future disease risk, *classification* as diagnostic support, or *generation* of various texts or images using natural language processing (NLP) in medicine [1-3].

NLP is an area of AI that addresses the interaction between human languages and machines [4]. The major roles of NLP in medicine and healthcare include serving as supportive tools in clinical practice and research [3]. Beyond the prediction of certain risk factors or clinical decision-making, NLP assists physicians and researchers to efficiently extract, translate, classify and analyze patients’ information and clinical-free text in electronic medical and health records, in addition to dialogue generation and answering medical information [3, 4]. The performance of NLP has dramatically improved following the emergence of transformer-based large language models (LLMs). A transformer is a type of neural network model that employs self-attention mechanism, relating multiple positions of a single sequence to compute a representation of the sequence [5]. LLMs are created using advanced ML techniques, especially deep neural networks, trained on enormous amounts of text data from the Internet and other sources [4]. A few notable LLMs include pretrained Bidirectional Encoder Representations from Transformers (BERT) [6], Language Models for Dialog Applications (LaMDA) [7], Pathway Language Model (PaLM) [8], Large Language Model Meta (LLaMA) [9], and Generative Pretrained Transformer (GPT)-3 and later models [10-12].

Recently, InstructGPT (GPT-3.5)—a GPT model employing 175 billion parameters with supervised fine-tuning and reinforcement learning from human feedback [11]—and its dialogue-optimized chatbot (ChatGPT) launched in November 2022 have significantly impacted NLP fields [13]. By predicting the subsequent element of the texts, ChatGPT can comprehend user prompts and generate human-like responses, expressed in ethical, sentimental, logical, and creative manner, without any additional training (*e*.*g*., foundation model) [14]. Although GPT is a non-domain-specific LLM, not exclusively intended to be used for medical or healthcare fields, recent publications have demonstrated that ChatGPT (GPT-3.5) possesses sufficient ability to pass the United States Medical Licensing Examination [15, 16]. In contrast, another study reported ChatGPT’s inadequate performance on non-English-based Korean medical questions [17]. Although the performance variation can be attributed to differences in languages, domestic healthcare systems, diagnostic criteria, and treatment strategies, the relationship between these differences and ChatGPT’s performance in answering medical questions remains unclear. Furthermore, the performance of ChatGPT with the current GPT-4 model employing an estimated 10 trillion parameters [12] has not yet been evaluated on the latest Medical Licensing Examination, which was originally written in non-English texts and held after the completion of GPT-4 model training (August 2022) [18].

Therefore, this study tested the performance of GPT (both GPT-3.5 and GPT-4 models) on the 117^th^ National Medical Licensing Examination (NMLE) (held in February 2023 in Japan), which was originally conducted in the Japanese language. In particular, questions from the previous year (116^th^ NMLE exam held in February 2022) were used as a model and prompt performance tuning set before using the latest questions (117^th^ exam held in February 2023) as a performance testing set to verify whether GPT can qualify for the actual minimal passing rate of this examination.

## Results

### Improving performance through English translation and tuned prompts in 116^th^ NMLE (2022)

Initially, we used the non-image-based questions from 116^th^ NMLE in Japan to develop the optimal input prompts for ChatGPT to maximize the correct answer rate. We extracted the question data from the 116th NMLE containing 394 questions (originally 400 questions, but six were officially removed from scoring evaluation). Thereafter, we removed questions with image data (n = 104) and analyzed the remaining 290 questions without image data (**Figure 1**).

**Figure 1.**
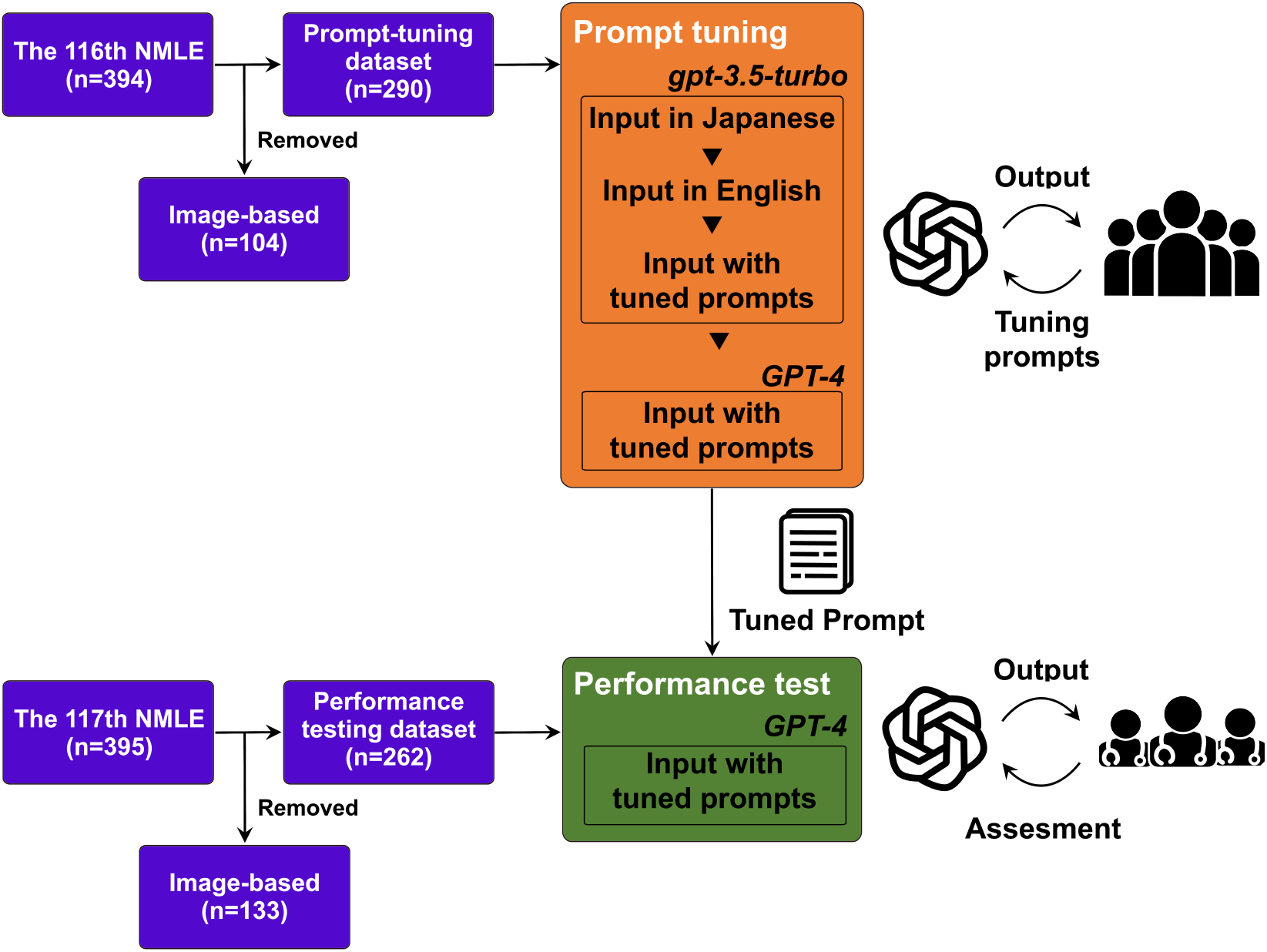
Study overview. Questions from the 116^th^ NMLE in Japan were used as the prompt-tuning dataset and those from 117^th^ NMLE were utilized as the performance-testing dataset after removing the image-based questions. During the prompt tuning process, questions from the prompt-tuning dataset were input into GPT-3.5-turbo and GPT-4, using simple prompts in both Japanese and English along with tuned prompts in English. Subsequently, we evaluated the outputs from GPT-3.5-turbo and GPT-4 with tuned prompts. After tuning the prompts, the ChatGPT (GPT-4) model optimized with the tuned prompts was tested on the performance-testing dataset (117^th^ NMLE).

Using the ChatGPT API powered by GPT3.5, we initially tested its performance for the original questions in Japanese language. Initially, we obtained a correct answer rate of 52.8% (153/290) with an output error rate of 5.5% (16/290). Accordingly, we used updated prompts to translate the original Japanese NMLE questions into English using ChatGPT before inputting them as questions. Although this marginally increased the correct answer rate to 56.2% (163/290), the output errors increased to 14.8% (43/290; **Figure 2**).

**Figure 2.**
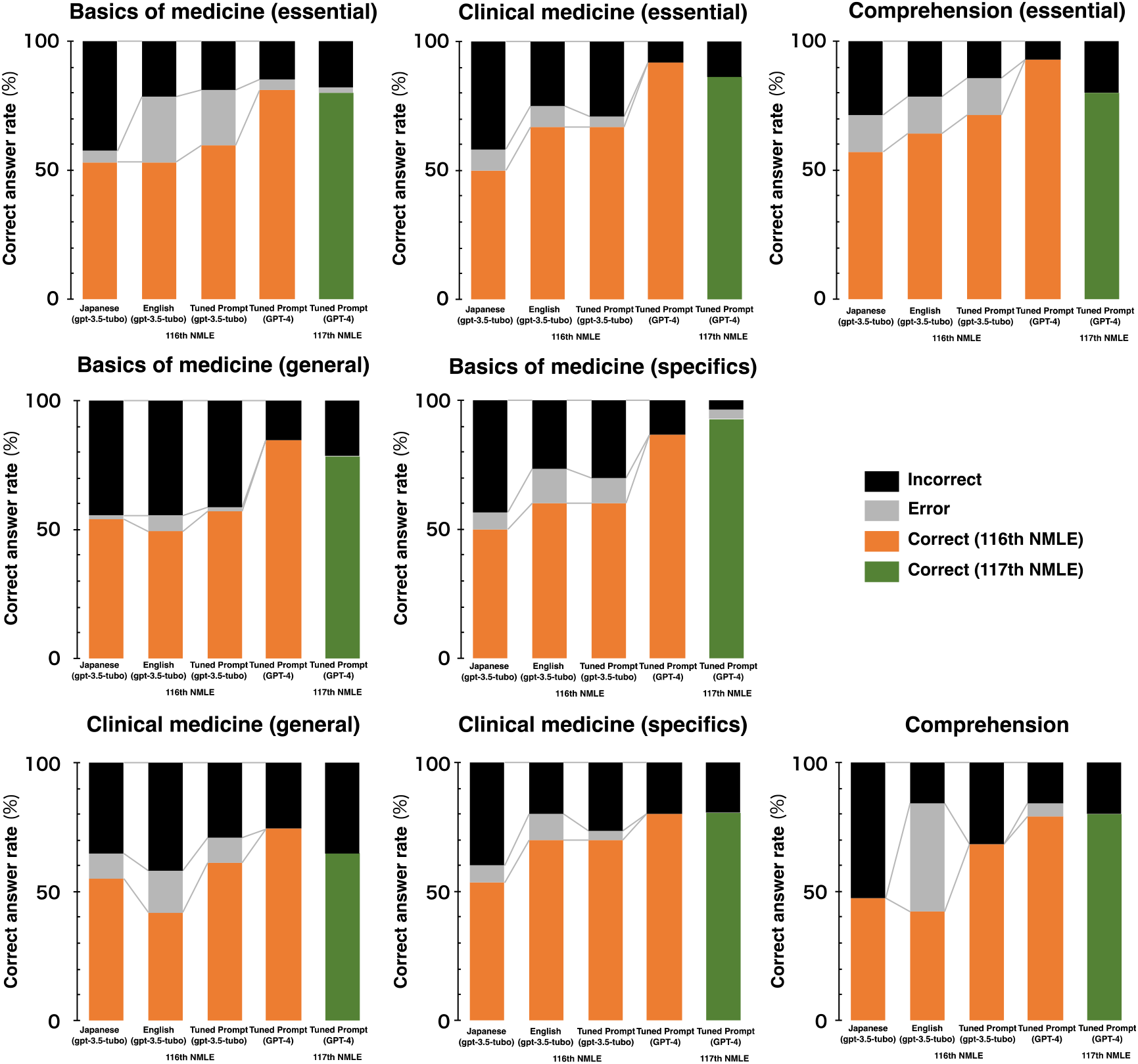
Variations in the rate of correct answers across languages, prompt tuning levels, and GPT models. Translating the Japanese questions into English text improved the correct answer rate; however, it increased the output error rate. Upon further tuning the prompts, the correct answer rate improved and the output error decreased. Moreover, switching from the GPT-3.5 model to the GPT-4 model enhanced the correct answer rate and almost eliminated errors.

To further improve the correct answer rate and reduce the errors, we tuned our prompts for each question type (Basics of Medicine, Clinical Medicine, and Comprehension). In particular, we provided sample outputs and directed the model to translate the questions into plain English and create summaries before answering the questions (**Figure 3**). This tuned prompt improved the correct answer rate to 63.1% (183/290) with a reduced output error rate of 7.6% (22/290). Furthermore, we applied the above-tuned prompts to the GPT-4-based ChatGPT, which demonstrated a correct answer rate of 82.8% (240/290) and a minimal error rate of 1.0% (3/290) (**Figure 2**).

**Figure 3.**
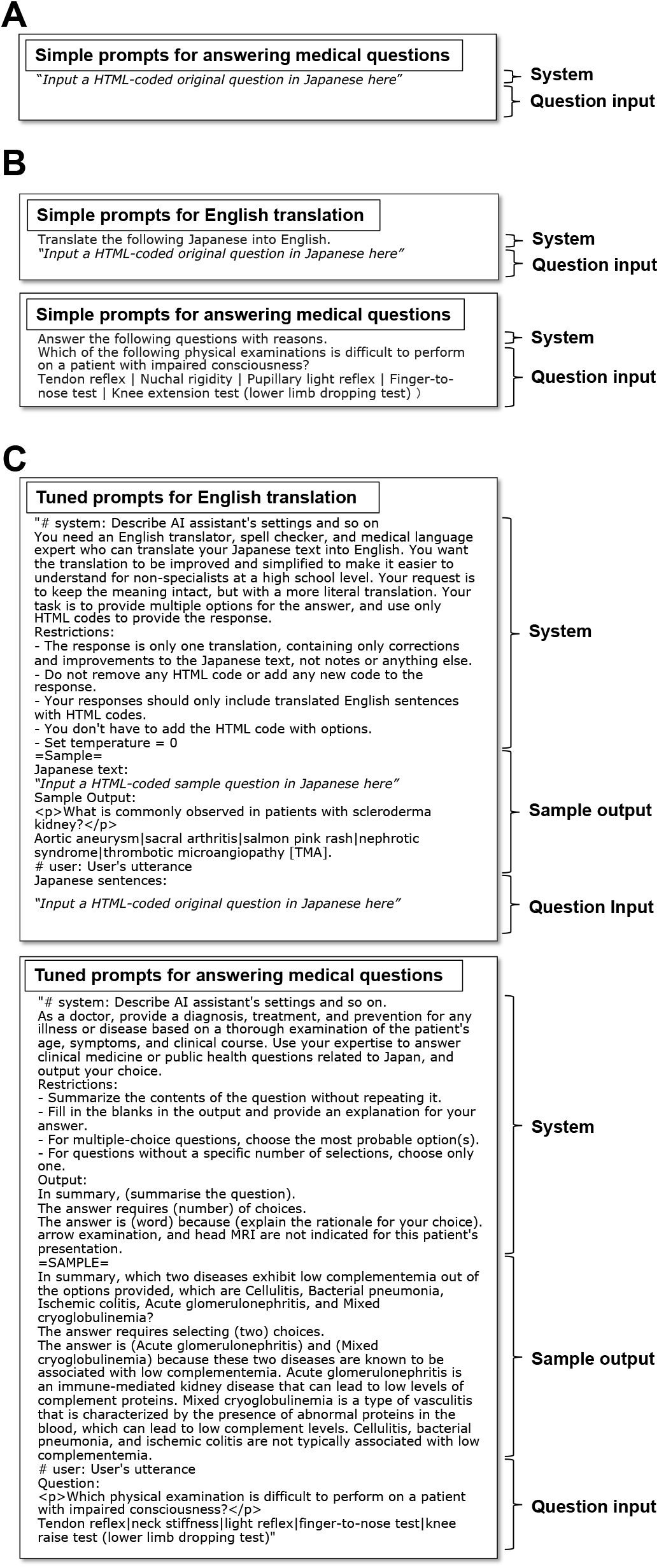
Examples of prompts for English translation and answering medical questions. **A:** A simple “Japanese prompt” used for answering Japanese questions. **B:** Simple “English prompts” used for Japanese-to-English translation and answering translated questions. **C:** Our optimized “English with tuned prompts”. The final optimized two-step prompts comprised a “system,” “sample output,” and “question input” sections. ChatGPT was initially instructed to translate HTML-based Japanese questions into simple, direct, and improved English. In both processes, the system of requirement and an exemplary output scenario were provided within the prompts. In the question input section, the Japanese questions were inputted to the English translation process, and sequentially, the English-translated questions were used to obtain the answers of the 117^th^ NMLE questions.

### GPT-4-based ChatGPT performance on 117^th^ (2023) NMLE with tuned prompt

Thereafter, we evaluated that the performance of the best model (GPT-4) with a tuned prompt for the test set of 262 questions without image data from the 117^th^ NMLE in Japan, held in February 4^th^ and 5^th^, 2023, after the completion of GPT-4 model training in August 2022 (**Figure 1**). With a tuned prompt, the best model achieved a correct answer rate of 78.6% (206/262) and an output error rate of 0.8% (2/262) (**Table 1**).

**Table 1.**
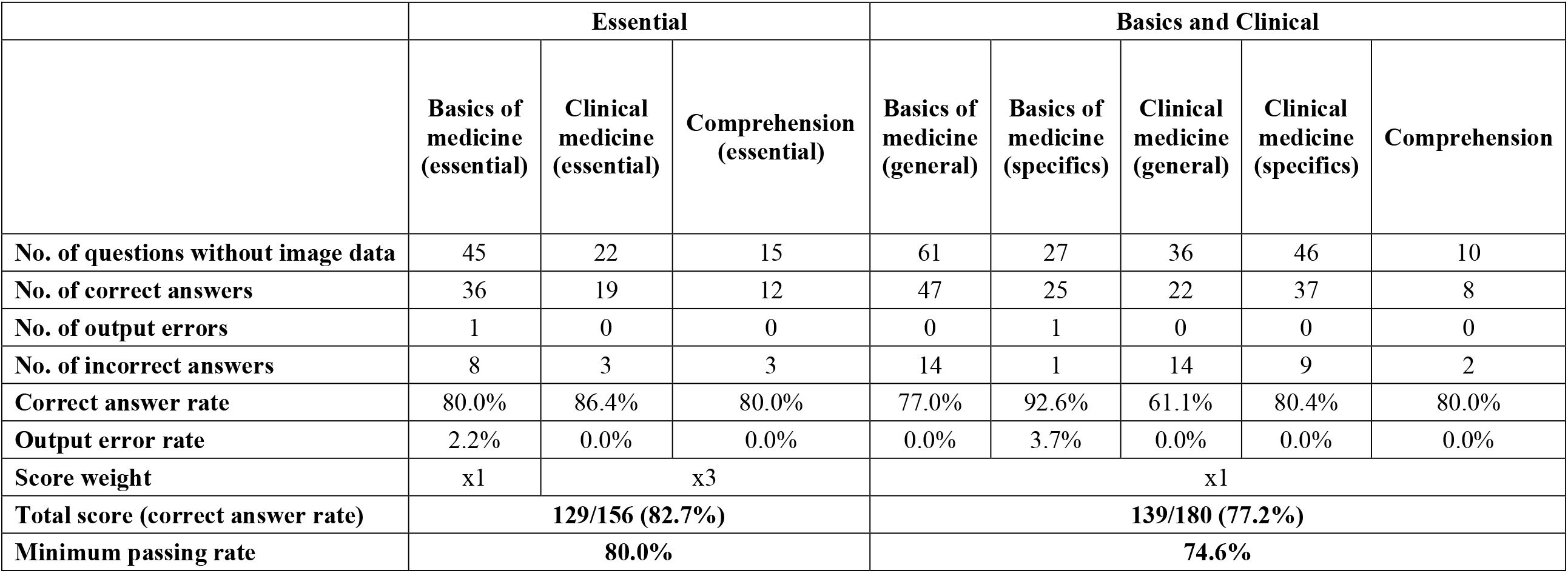
Performance of optimal GPT-4 model with tuned prompt for the 117^th^ NMLE in Japan.

|  | Essential |  |  | Basics and Clinical |  |  |  |  |
| --- | --- | --- | --- | --- | --- | --- | --- | --- |
|  | Basics of<br>medicine<br>(essential) | Clinical<br>medicine<br>(essential) | Comprehension<br>(essential) | Basics of<br>medicine<br>(general) | Basics of<br>medicine<br>(specifics) | Clinical<br>medicine<br>(general) | Clinical<br>medicine<br>(specifics) | Comprehension |
| No. of questions without image data | 45 | 22 | 15 | 61 | 27 | 36 | 46 | 10 |
| No. of correct answers | 36 | 19 | 12 | 47 | 25 | 22 | 37 | 8 |
| No. of output errors | 1 | 0 | 0 | 0 | 1 | 0 | 0 | 0 |
| No. of incorrect answers | 8 | 3 | 3 | 14 | 1 | 14 | 9 | 2 |
| Correct answer rate | 80.0% | 86.4% | 80.0% | 77.0% | 92.6% | 61.1% | 80.4% | 80.0% |
| Output error rate | 2.2% | 0.0% | 0.0% | 0.0% | 3.7% | 0.0% | 0.0% | 0.0% |
| Score weight | x1 | x3 |  | x1 |  |  |  |  |
| Total score (correct answer rate) | 129/156 (82.7%) |  |  | 139/180 (77.2%) |  |  |  |  |
| Minimum passing rate | 80.0% |  |  | 74.6% |  |  |  |  |

The present results were compared with the actual minimal passing rate on the examination. The current model with a tuned prompt scored 82.7% (129/156) for essential questions and 77.2% (139/180) for basic and clinical questions, both of which qualified the minimum passing rates of 80.0% and 74.6%, respectively (**Figure 2**) [19]. Notably, we applied the GPT-4 model with tuned prompts to the entire set of 395 questions (text-only) in the 117^th^ NMLE, regardless of containing image data (originally 400 questions, but five were officially removed from scoring evaluation). This optimal model attained near-passing levels of 78.5% (157/200) for essential questions and 73.2% (216/295) for basic and clinical questions.

### Exploratory analysis of incorrect ChatGPT responses and their associated explanations

To further enhance the performance of the model, we performed an exploratory analysis of 56 incorrect answers provided by the optimal GPT-4 model with tuned prompts for the 117^th^ NMLE questions. As listed in **Table 2**, the three primary factors contributing to the generation of incorrect answers by the model included insufficient medical knowledge (33/56, 58.9%), Japan-specific medical system information (17/56, 30.4%), and mathematical errors (4/56, 7.1%). Concerning the insufficient medical knowledge, the areas of incorrect answers were not specific and spanned across various medical fields. Notably, certain answers were outdated or critically incorrect in current medical contexts (**Figure 4**). In terms of Japan-specific medical system, ChatGPT failed to adequately answer questions related to Japanese medicolegal laws applicable in the medical and healthcare field, guidance from the Ministry of Health, Labour, and Welfare (MHLW) in Japan, and guidelines, especially those related to public health. Additionally, we noted several mathematical errors such as in addition calculations (*e*.*g*., the explanation and addition formula were correct, but the answer was wrong) and handling decimal points (because of translation errors from the phrase “rounding to first decimal point” from Japanese).

**Table 2.** Summary of incorrect answers from the optimal model.

| **Total incorrect answer** | **N = 56** |
| --- | --- |
| **Insufficient medical knowledge** | **33 (58.9%)** |
| Breast surgery | 1 |
| Dermatology | 2 |
| Emergency medicine | 2 |
| Endocrinology | 6 |
| Gastroenterology | 2 |
| Immunology | 1 |
| Medical interview | 1 |
| Medical procedure | 1 |
| Nephrology | 2 |
| Neurology | 1 |
| Obstetrics and gynecology | 2 |
| Ophthalmology | 1 |
| Pediatrics | 2 |
| Physical examination | 1 |
| Psychiatry | 1 |
| Public health | 1 |
| Rehabilitation | 1 |
| Respiratory medicine | 3 |
| Rheumatology | 1 |
| Urology | 1 |
| **Japan-specific medical system** | **17 (30.4%)** |
| Clinical research | 1 |
| Emergency | 1 |
| Psychiatry | 1 |
| Public health | 14 |
| **Mathematical errors** | **4 (7.1%)** |
| Respiratory | 1 |
| Pediatrics | 1 |
| Cardiology | 1 |
| Medical interview | 1 |
| **Others** | **2 (3.6%)** |
| Issue in English translation | 1 |
| Not providing an answer | 1 |

**Figure 4.**
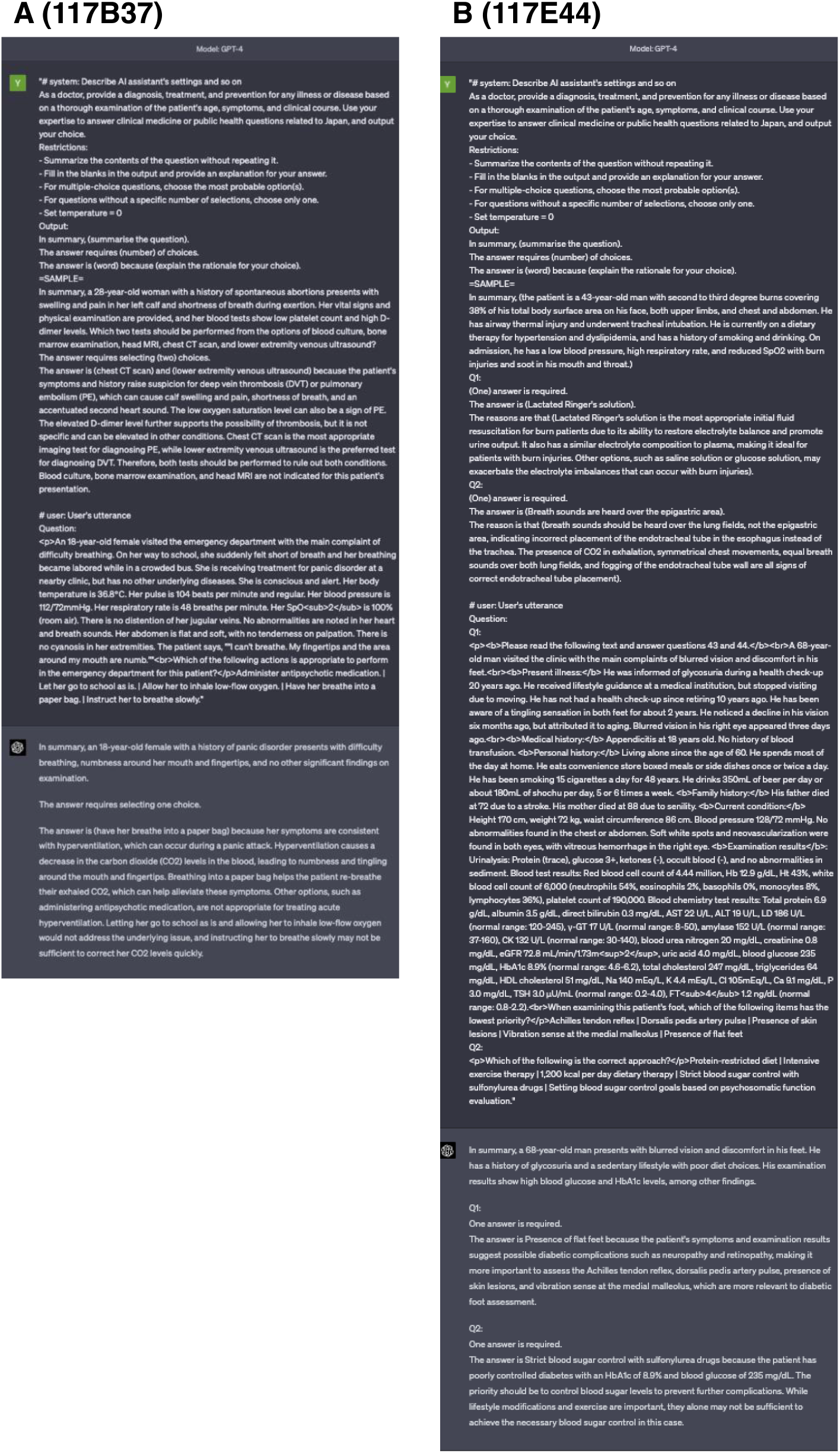
Examples of potentially outdated or critically incorrect outputs from the model in current medical contexts. **A:** A question on the primary treatment for hyperventilation syndrome in the emergency department. The suggestion of paper bag method for raising the carbon-dioxide concentration in the blood has been commonly used in the past, but it is not always the first choice, as it can worsen symptoms in certain patients with secondary hyperventilation, *e*.*g*., those with lung diseases causing low blood oxygen levels. In such answers, it seems that outdated, traditional information can prevail over the latest information, especially if it has been a standard practice over a period and related information is widely available on the Internet. **B:** A question on the initial outpatient treatment for a type-2 diabetes patient with poor control and combined diabetic retinopathy and neuropathy. The long-term treatment goal for diabetes is strict blood sugar control, but in this case, strict blood sugar control with sulfonylurea drugs during the initial treatment may aggravate the risk of diabetic retinopathy, raising strong concerns on ChatGPT’s answer.

## Discussion

This study evaluated the performance of GPT on the Japanese Medical Licensing Examination. The results indicate that 1) GPT-4 with a tuned prompt cleared the minimal passing rate on the 116^th^ (2022) NMLE in Japan; 2) GPT-4 with tuned prompt qualified the minimum passing rate on the latest 117^th^ NMLE (2023); and 3) Inadequate medical knowledge, Japan-specific medical system information, and mathematical errors were the primary factors associated with the incorrect answers generated by the optimal model. Despite the absence of image data in the questions, this study demonstrated the first attempt to use the best available ChatGPT model with tuned prompts to achieve a minimum passing rate for the latest 117^th^ NMLE in Japan.

This study provides several conclusions. First, GPT-4 with a tuned prompt cleared the minimal passing rate on the 116^th^ NMLE in Japan held in February 2022. Although GPT-3.5-based ChatGPT achieved a correct answer rate of 52.8% for Japanese questions, it increased to 56.2% after translating the questions into English. As GPT-3, the original GPT-3.5, was primarily trained in English, it delivers a higher performance when responding to prompts in English compared to other languages [10]. Similarly, a recent multilingual performance evaluation of GPT-4, an improved version of GPT-3, confirmed that the best performance is more generally obtained with English prompts [12]. After tuning our prompts to include a translation procedure into plain English and modifying the output format based on the question type, the correct response rate increased to 63.1%. This finding is consistent with previous studies claiming that prompt engineering can improve model task performance [12, 20]. These improved correct response rates can be attributed to English being the majority of the language in the training data, *i*.*e*., the Internet, used by non-experts [21]. Although the error rate increased to 14.8% upon translating the Japanese questions into English, it notably decreased to 7.6% after tuning the prompts by including the format of the output. This result suggests that providing samples and standardizing the output format can produce the desired output format and reduce the number of errors. Finally, upon applying these optimized prompts to GPT-4-based ChatGPT, the correct response rate increased to 82.8% and the error rate plummeted to 1.0%. This significant improvement in performance can be ascribed to the advanced architecture and training of GPT-4 [12].

Second, even in case of the latest 117^th^ NMLE (2023), GPT-4 with tuned prompt qualified the actual minimum passing rate. GPT-4 has passed various professional examinations in English, including the practice bar exam with a score in the top 10% of examinees [12]. A previous study reported that ChatGPT (GPT-3.5) failed to achieve the minimum passing rates [22]. However, this study demonstrated that ChatGPT (GPT-4) can pass the 117^th^ NMLE with the optimized prompt tuning method proposed herein. The current results can be derived from the exquisite combination of essential factors such as English translation and optimally tuned prompts for obtaining correct answers through the best performance of the latest ChatGPT model.

Third, inadequate medical knowledge, information related to the medical and healthcare system guidelines of Japan, and mathematical errors formed the three major factors of the incorrect answers generated by the best available ChatGPT model with tuned prompts. Among the incorrect answers associated under inadequate medical knowledge, no significant bias was observed for the medical fields relevant to each question. Furthermore, even after providing incorrect answers, the model output plausible but wrong medical explanations (so-called hallucinations in LLM outputs [23]). Therefore, even if the model exhibits a performance level that surpasses the minimum score for the NMLE, a broader range of specialized and up-to-date medical knowledge regarding standard treatments should be inputted. In addition, output receivers should be equipped with professional medical knowledge to assess the correctness of the output. For the Japan-specific system, several incorrect answers were observed, especially in public health-related questions, which are based on Japanese laws, guidelines, and unique systems. Although the GPT-4 powered ChatGPT delivered improved performance in terms of output differences between the languages, every country should perform their individual localization in terms of the applicable laws and systems considering the language differences. Furthermore, in certain cases related to mathematical errors, the calculation formula in the explanation was correct, but the result and the final answer output were incorrect. Moreover, an instruction of “approximating the decimal place” was not properly comprehended by ChatGPT during the Japanese-to-English translation. As such, calculation problems are reported as one of the areas where LLMs still exhibit relatively low accuracy [24], indicating that calculation problems may be a relatively unsuitable field for current ChatGPT.

As discussed, we express strong concerns regarding the use of the current ChatGPT for medical purposes, as OpenAI has already indicated that the models should not be used for providing triage, diagnosis, or treatment options for life-threating issues or severe medical conditions [25]. Indeed, for use in medical settings, an approval must be obtained from regulatory agencies, *e*.*g*., software as a medical device. Moreover, utilizing such technology is already difficult with its several black-box aspects [12]. Various countries have released statements regarding the applications of LLMs in medical fields [26, 27]. Although the versatility of these models hinders the verification of their validity and they require enormous computational resources and costs, we believe that the advanced medical foundation AI model [28] can replace task-specific approach AI models and will appear not far off, with scientifically proven clinical efficacy and safety in medical and healthcare fields.

The novelty of this study is that it is the first research to achieve a minimum passing rate using 262 non-image questions in the latest 117^th^ NMLE in Japan with the ChatGPT GPT-4 version with the optimally tuned prompts. The limitations of this study were as follows. First, we only used questions without image data to evaluate the performance of the best available model with tuned prompts, although it might be fair to assess the ability of the model to pass the examination using all questions, regardless of image data. However, as revealed from the Results, we observed a favorable model performance even upon using the entire question set in the 117^th^ NMLE in Japan. Second, the NMLE in Japan uniquely included strongly not-recommended “contraindication” answer choices within the questions. The MHLW in Japan has set the minimum passing criteria regarding selecting contraindication answer choices to be equal or less than three for the 116^th^ NMLE or two for the 117^th^ NMLE. As the real number of contraindication answer choices were not officially announced by the MHLW, we could not use them in the current performance evaluation.

In conclusion, GPT-4 powered ChatGPT with optimally tuned prompts achieved a minimum passing rate in the latest 117^th^ NMLE in Japan. In addition, the model scored near-passing levels for the entire test dataset of 395 questions, regardless of medical image data. The upcoming GPT-4 version, which features enhanced image recognition capabilities, will easily qualify the minimum passing rate and achieve top-tier scores, as reported in other English-based examinations [12]. We again express strong concerns in terms of using of the current ChatGPT for medical purposes so far. However, beyond its original design of answering examination questions for humans, these AI models might have the potential be regarded as one of the best “sidekicks” for solving problems and fulfilling the current needs in the medical and healthcare fields in the near future.

## Materials and methods

### Study overview

This study evaluated the performance of GPT models on the NMLE in Japan. We utilized both the GPT-3.5 and GPT-4 models of ChatGPT (Open AI, Inc., San Francisco, CA, USA). Initially, the questions from the 116^th^ NMLE in Japan (February 2022) were used as a model and prompt tuning set to optimize the performance of obtaining the correct answers and explanations. Subsequently, we assessed the performance of the best ChatGPT model (GPT-4) with the tuned prompts for answering the questions from the 117^th^ NMLE in Japan (February 2023).

### Input source

The questions and answers for the 116^th^ NMLE in Japan were obtained from the official website of the MHLW, Japan [29]. For the latest 117^th^ NMLE, we manually performed optical character recognition on the original question papers to create input data and extracted the official answers from the MHLW website [19]. The examination comprised six blocks (A–F), with 75 questions in blocks A, C, D, and E, and 50 questions in blocks B and F. Note that six questions in the 116^th^ NMLE and five in the 117^th^ NMLE were excluded. In addition, all image-containing questions were removed from both the prompt-tuning and the performance-testing datasets, because up till early April 2023, only text-based questions could be used as input to the ChatGPT interface, including the API. The number of image-containing questions was 104 in the 116^th^ NMLE and 133 in the 117^th^ NMLE. Thereafter, according to the Japanese NMLE scoring method, the remaining questions without image data were classified into the categories of “Essential” and “Basic and Clinical.” The 116^th^ NMLE in Japan included 47 questions related to basics of medicine (essential), 24 questions of clinical medicine (essential), 14 questions on comprehension (essential), 65 questions regarding basics of medicine (general), 30 questions in basics of medicine (specifics), 31 questions of clinical medicine (general), 60 questions of clinical medicine (specifics), and 19 questions on comprehension. The 117^th^ NMLE in Japan comprised 45 questions related to basics of medicine (essential), 22 questions of clinical medicine (essential), 15 questions on comprehension (essential), 61 questions from the basics of medicine (general), 27 questions on the basics of medicine (specifics), 36 of clinical medicine (general), 46 questions related to clinical medicine (specifics), and 10 questions regarding comprehension. Finally, we used 290 questions (without image data) from the 116^th^ NMLE and 262 questions (without image data) from the 117^th^ NMLE in Japan for analyses. The entire set of 395 text-based questions, irrespective of image data, from the 117^th^ NMLE in Japan was considered for the exploratory analysis.

### Generative Pretrained Transformer

The GPT, developed by OpenAI [14], is a type of AI model used for NLP tasks. Following the research path from the original GPT, GPT-2, and GPT-3, OpenAI’s DL approach leverages extensive amounts of data and intensive computation to create increasingly sophisticated and capable language models [18]. ChatGPT has been fine-tuned from the initial GPT-3.5, and later, GPT-4—a LLM trained in early 2022 to produce text [13, 30]. GPT-4 is OpenAI’s latest and most advanced AI model that can solve difficult problems with greater accuracy [18]. In this study, we used ChatGPT powered by both the GPT-3.5 and GPT-4 versions.

### Prompt engineering to maximize the correct answer rate

We used the 116^th^ NMLE in Japan to generate the most suitable prompts for ChatGPT to answer the 117^th^ NMLE questions. Using the ChatGPT API, we first instructed ChatGPT to respond to the original questions in Japanese language. We manually coded the Hyper Text Markup Language (HTML) to represent the bold, italic, superscript, and subscript characters in the original text (**Figure 3A**). Second, we instructed ChatGPT to translate the original Japanese NMLE questions into English using its own capabilities before inputting them as questions (**Figure 3B**). In addition, we compiled and analyzed the output errors. Thereafter, we provided prompts with restriction sentences designed to prevent the reoccurrence of these errors, along with sample outputs illustrating the desired output format. Finally, we inquired ChatGPT to improve the prompt itself. We further refined the prompts using the 116^th^ NMLE questions to achieve higher rates of correct answers and output in the desired format, because prompt tuning can improve the task accuracy compared to training the entire model [12, 20]. The final optimized two-step prompts for the English translation process and the process of answering the medical questions are illustrated in **Figure 3C**, wherein each process comprised “system,” “sample output,” and “question input” sections. We organized the output examples according to each medical question category (basics of medicine, clinical medicine, and comprehension). In brief, ChatGPT was initially instructed to translate the HTML-based Japanese questions into plain, direct, and improved English, while maintaining the original HTML codes without deleting or adding new text. In both processes, the system of requirement and an exemplary output scenario were provided within the prompts. In the question input section, the HTML-based Japanese questions were inputted for the English translation process, and the English-translated questions were consequently inputted to the process of answering the medical questions (**Figure 3)**. To minimize output variability, all input prompts were executed with the temperature parameter set to 0.

GPT-3.5-based analyses were performed using the ChatGPT API with custom Python code on the Google Colaboratory interface. GPT-4-based analysis was conducted using ChatGPT website console, with eight investigators (Y. T., T. N., K. A., T. E., R. M., S. K., H. K., and F. H.) manually inputting prompts one by one and changing a thread each time. Specifically, they inputted the questions, choices, and appropriate prompts into ChatGPT and summarized the output answers. We used the GPT-3.5 version GPT3.5-turbo-0301 for the “Japanese,” “English,” and “English with tuned prompt” analyses, and the GPT-4 model version released on March 14^th^ 2023 for the “English with tuned prompt” analysis.

### Outcomes

The target outcome of this study is the correct answer rate. We manually compared ChatGPT’s output answers with the official answers to determine the correctness of the output answers. Accordingly, the correct answer rate was calculated as the number of correct answers divided by the number of questions. We defined the output errors as incorrect answers. To evaluate the potential performance for passing the 117^th^ NMLE in Japan, we applied the minimum passing rates, not the minimum passing scores, to evaluate the model performance because the image-containing questions were excluded from the analyses.

### Performance evaluation

In the primary performance evaluation, we assessed the correct answer rate for questions without images in the 117^th^ NMLE in Japan using the best ChatGPT model (GPT-4) with tuned prompt, which was compared to the actual minimally passing rate on the examination. In the secondary performance evaluation, we examined the correct answer rate for all questions in the 117^th^ exam using the optimal model and prompts. In addition, the medical reasonableness of the generated explanations for each answer was assessed by two independent clinical physicians (M.N. and M.K.) and was double-checked by another independent clinical physician (A.N.). Furthermore, we analyzed the content of the incorrect answers along with their explanations to identify the areas in which the application of the current ChatGPT for medicine may be relatively weak.

## Data Availability

The whole input questions and answers from the model for the 117th NMLE in Japan are listed in the Supplemental Data. The ChatGPT APIs used in this study are accessible via GitHub (https://github.com/yudaitanaka1026/ChatGPT_NMLE_Japan).

## Acknowledgments

We express our gratitude to Yasuhiro Onogi and Yuichi Miyamae at MICIN, Inc. for their insightful online discussions regarding this project. We thank Dr. Hozumi for dedicating his time to discuss this topic with us. We also thank ChatGPT (GPT-4) and Enago English proofreading service for English proofreading.

## Data availability

The ChatGPT APIs used in this study are accessible via GitHub (https://github.com/yudaitanaka1026/ChatGPT_NMLE_Japan).

## Conflict of Interest

The authors declare no conflicts of interest relevant to this article.

## Financial disclosure

None.

## Author contributions

**Conceptualization:** Yudai Tanaka, Takuto Nakata, Ko Aiga, Hiroaki Kakizaki, and Akihiro Nomura.

**Data curation:** Yudai Tanaka, Takuto Nakata, Ko Aiga, Takahide Etani, Ryota Muramatsu, Shun Katagiri, Hiroyuki Kawai, Fumiya Higashino, and Masahiro Enomoto.

**Formal analysis:** Yudai Tanaka.

**Methodology:** Yudai Tanaka, Takuto Nakata, Hiroaki Kakizaki, and Akihiro Nomura.

**Project administration:** Akihiro Nomura.

**Supervision:** Masayuki Takamura, Takashi Yoneda, and Hiroaki Kakizaki.

**Validation:** Masao Noda, Mitsuhiro Kometani, and Akihiro Nomura.

**Visualization:** Yudai Tanaka, Takuto Nakata, Ko Aiga, and Akihiro Nomura.

**Writing – original draft:** Yudai Tanaka, Takuto Nakata, Ko Aiga, and Akihiro Nomura.

**Writing – review and editing:** Yudai Tanaka, Takuto Nakata, Ko Aiga, Takahide Etani, Ryota Muramatsu, Shun Katagiri, Hiroyuki Kawai, Fumiya Higashino, Masahiro Enomoto, Masao Noda, Masayuki Takamura, Mitsuhiro Kometani, Takashi Yoneda, Hiroaki Kakizaki, and Akihiro Nomura.

## Notes

### Competing Interest Statement

The authors have declared no competing interest.

